# Machine Learning-driven Histotype Diagnosis of Ovarian Carcinoma: Insights from the OCEAN AI Challenge

**DOI:** 10.1101/2024.04.19.24306099

**Authors:** Maryam Asadi-Aghbolaghi, Hossein Farahani, Allen Zhang, Ardalan Akbari, Sirim Kim, Ashley Chow, Sohier Dane, OCEAN Challenge Consortium, OTTA Consortium, David G Huntsman, C Blake Gilks, Susan Ramus, Martin Köbel, Anthony N Karnezis, Ali Bashashati

**Author notes:** Corresponding author Correspondence: Ali Bashashati. Mailing address: 2222 Health Sciences 25 Mall, Vancouver, BC V6T 1Z3 Canada. Joint senior authors (Anthony N Karnezis, Ali Bashashati). These authors contributed equally to this work (Maryam Asadi-Aghbolaghi, Hossein Farahani, Allen Zhang).

## Abstract

Ovarian cancer poses a significant health burden as one of the deadliest malignancies affecting women globally. Histotype assignment of epithelial ovarian cancers can be challenging due to morphologic overlap, inter-observer variability, and the lack of ancillary diagnostic techniques in some areas of the world. Moreover, rare cancers can pose particular diagnostic difficulties because of a relative lack of familiarity with them, underscoring the necessity for robust diagnostic methodologies. The emergence of Artificial Intelligence (AI) has brought promising prospects to the realm of ovarian cancer diagnosis. While various studies have underscored AI’s promise, its validation across multiple healthcare centers and hospitals has been limited. Inspired by innovations in medical imaging driven by public competitions, we initiated the Ovarian Cancer subtypE clAssification and outlier detectioN (OCEAN) challenge — the most extensive histopathology competition to date.

## Introduction

Epithelial ovarian cancers are some of the most lethal gynecologic malignancies in North America and across the world [1]. This disease exhibits marked heterogeneity, characterized by five major histotypes: high-grade serous carcinoma (HGSC), constituting 70% of cases (and 90% of advanced-stage disease and mortality); clear cell ovarian carcinoma (CCOC), representing 12%; endometrioid (ENOC) at 11%; low-grade serous (LGSC) at 4%; and mucinous carcinoma (MUC) at 3% [2, 3], and several additional rare subtypes histotypes. Clinical management varies greatly between histotypes, with high-grade serous carcinomas treated most aggressively with combination platinum-taxane chemotherapy and a subset benefitting from PARP inhibitor therapy and MEK inhibitors [4]. Endometrioid and clear cell carcinomas are associated with mismatch repair deficiency, which can be sporadic or inherited due to Lynch syndrome. Accurate diagnosis histotyping is critical for risk assessment, hereditary cancer screening, and clinical trial enrollment. However, histological classification of ovarian carcinomas by pathologists still suffers from suboptimal interobserver and intraobserver reproducibility outside of gynecologic pathology subspecialty practice and tertiary academic centers [3, 5–8].

Initial diagnosis relies on histological examination of hematoxylin and eosin (H&E)-stained tissue sections, yet studies indicate that among pathologists lacking specialty training in gynecologic pathology, the interobserver agreement in diagnosis remains only moderate (0.54–0.67 Cohen’s kappa) [9, 10]. While adjunct diagnostic techniques such as immunohistochemistry and next-generation sequencing can improve diagnostic accuracy and interobserver reproducibility [7, 10–12], these techniques are unavailable in much of the world [13, 14]. Additionally, there is a pronounced shortage of pathologists compared to the demand, leading to substantial variances in pathologist numbers among countries [15]. Even in well-equipped nations, the demand for pathologists surpasses the available supply [14, 16]. Addressing these challenges is imperative to ensure effective diagnosis and management of ovarian carcinoma patients [17].

In recent years, the integration of AI algorithms into the field of medical diagnostics has shown considerable promise [18–23], particularly in aiding pathologists with the histological classification of ovarian cancer [24, 25]. However, despite their potential, AI algorithms are not immune to biases that may arise during their development and validation processes. These biases can manifest in algorithms that perform suboptimally when applied to datasets beyond those used in their initial training, highlighting the critical need for robust methodologies to assess their generalizability. To facilitate this endeavor, we launched the Ovarian Cancer subtypE clAssification and outlier detectioN (OCEAN) challenge, a global competition that provided participants with access to the largest and most diverse public histopathology dataset of ovarian cancer to date.

The OCEAN dataset comprises approximately 2500 samples, encompassing both whole slide images and tissue microarrays sourced from over 20 centers across multiple countries. Variations in patient demographics, tissue processing, and H&E slide staining protocols across pathology labs contribute to diversity in the dataset. The extensive variation in color among H&E slide samples presents a unique opportunity for assessing the generalizability and robustness of algorithms versus those trained on slides from a single center [25]. Importantly, since the evaluation is conducted independently of algorithm development, we mitigate the risk of information leakage.

Through the OCEAN challenge, our dedication lies in expediting progress and establishing the foundation for developing AI solutions that make tangible clinical differences in diagnosing and managing ovarian cancer. By making this expansive dataset as well as top-performing AI models publicly accessible, our goal is to provide a significant resource for advancing research in ovarian cancer diagnosis and treatment, ultimately leading to improved patient outcomes, in particular for resource-limited practice settings.

## Results

### Dataset

The OCEAN dataset comprises 2,438 images distributed across three distinct sets: training, public test, and private test, sourced from 24 centers that were mainly a part of the ovarian tumor tissue analysis (OTTA) consortium [26]. This dataset encompasses H&E images from both Whole Slide Images (WSIs) and Tissue Microarrays (TMAs). Specifically, 538 images are allocated to the training set, while 437 and 1,488 images are designated for the public and private test sets, respectively. Within these images, the five main subtypes of ovarian carcinoma are represented: CCOC, ENOC, HGSC, LGSC, and MUC. Notably, the public and private test sets collectively contain 147 outlier images (i.e., other), comprising rare ovarian cancer subtypes along with normal tissues. For further details regarding the composition of the OCEAN dataset, please refer to Table 1.

**Table 1:** Overview of the OCEAN dataset.

| Set | Subtype | Train | Public Test | Private Test |
| --- | --- | --- | --- | --- |
| TMAs | CCOC | 5 | 64 | 175 |
|  | ENOC | 5 | 41 | 210 |
|  | HGSC | 5 | 51 | 492 |
|  | LGSC | 5 | 36 | 212 |
|  | MUC | 5 | 20 | 56 |
|  | Other | 5 | 31 | 44 |
| WSIs | CCOC | 94 | 23 | 61 |
|  | ENOC | 119 | 35 | 70 |
|  | HGSC | 217 | 68 | 71 |
|  | LGSC | 42 | 10 | 39 |
|  | MUC | 41 | 9 | 35 |
|  | Other | 0 | 49 | 23 |
| Total | All | 538 | 437 | 1,488 |

### OCEAN Competition

The OCEAN competition commenced on October 6th, 2023, and concluded on January 3rd, 2024, spanning a duration of three months. This competition was hosted on the Kaggle platform, a venue for data science challenges and collaborations. The event garnered significant interest, amassing a total of 9,247 registrations from participants worldwide. A total of 1,772 individuals comprising 1,326 teams representing 84 countries actively engaged in the competition. Throughout the duration of the competition, participants collectively submitted 35,279 entries in pursuit of refining their algorithms. Throughout the competition, participants had the chance to evaluate the effectiveness of their algorithms using the publicly provided test set. Notably, the leaderboard standings were determined based on the performance of algorithms on this public test set. Subsequently, winners were selected based on their algorithms’ performance on a separate private test set after the competition closing, ensuring a fair and unbiased evaluation process.

### Performance on Public and Private Datasets

Balanced accuracy served as the primary metric for assessing the efficacy of the submitted methodologies. Balanced accuracy, calculated as the average recall across all classes, addresses the issue of class imbalance by providing a comprehensive assessment of performance. Participants were required to assign each image to one of the five ovarian cancer histotypes or designate it as an “other” category within their submission files. Among the top 10 performing submissions on the public test set, balanced accuracy ranged from 61% to 68%. On the private test set, the top 10 performances exhibited balanced accuracies spanning from 58% to 66%.

## Conclusion and Discussion

The OCEAN challenge represents a significant step forward in the pursuit of accurate and robust AI solutions for the classification of ovarian carcinoma histotypes and the detection of outliers. The competition, hosted on the Kaggle platform, attracted substantial global participation, highlighting the widespread interest and commitment to advancing medical diagnostics through AI technologies.

The OCEAN dataset stands as a milestone in the field, emerging as the largest competition on histopathology images to date. It offers a comprehensive collection of histopathology images of ovarian carcinoma, unparalleled in its size and diversity. By subjecting algorithms to rigorous evaluation across datasets sourced from numerous hospitals, the OCEAN challenge sought to fill a crucial gap in the field. Specifically, it aimed to develop methodologies capable of robust generalization across diverse patient demographics, and digital slide scanners, tissue processing, and staining protocols across pathology labs.

Moving forward, the insights gained from the OCEAN challenge serve as a valuable foundation for further research and development efforts aimed at refining AI algorithms for the accurate diagnosis and classification of ovarian carcinoma histotypes.

## Data Availability

Data can be accessed and downloaded from the Kaggle challenge page (https://www.kaggle.com/competitions/UBC-OCEAN).

## Acknowledgements

This work was supported by BC Cancer Foundation, CIHR (No 418734), NSERC (RGPIN-2019-04896), and Health Research BC grants to AB.

## Notes

### Competing Interest Statement

The authors have declared no competing interest.

### Funding Statement

This work was supported by BC Cancer Foundation, CIHR (No 418734), NSERC (RGPIN-2019-04896), and Health Research BC grants to Dr. Ali Bashashati.

### Author Declarations

The Declaration of Helsinki and the International Ethical Guidelines for Biomedical Research Involving Human Subjects were strictly adhered throughout the course of this study. All study protocols have been approved by the University of British Columbia/BC Cancer Research Ethics Board.

